# Enhancing the sensitivity of rapid antigen detection test (RADT) of different SARS-CoV-2 variants and lineages using fluorescence-labeled antibodies and a fluorescent meter

**DOI:** 10.1101/2022.12.04.22283067

**Authors:** Gheyath K. Nasrallah, Fatma Ali, Salma Younes, Heba A. Al-Khatib, Asmaa A. Al-Thani, Hadi M. Yassine

**Affiliations:** Biomedical Research Center, Qatar University, Doha P.O. Box 2713, Qatar; Biomedical Sciences Department, College of Health Sciences, Qatar University, Doha P.O. Box 2713, Qatar

**Keywords:** RADT, SARS-CoV-2, COVID-19, Lateral flow, immunofluorescence

## Abstract

RT-qPCR is considered the gold standard for diagnosis of COVID-19; however, it is laborious, time-consuming, and expensive. RADTs have evolved recently as relatively inexpensive methods to address these shortcomings, but their performance for detecting different SARS-COV-2 variants remains limited. RADT test performance could be enhanced using different antibody labeling and signal detection techniques. Here, we aimed to evaluate the performance of two Wondfo antigen RADTs for detecting different SARS-CoV-2 variants: (i) the conventional colorimetric RADT (Ab-conjugated with gold beads); and (ii) the new Finecare™ RADT (Ab-coated fluorescent beads). Finecare™ is a meter used for the detection of a fluorescent signal. 187 frozen nasopharyngeal swabs collected in Universal transport (UTM) that are RT-qPCR positive for different SARS-CoV-2 variants were selected, including 60 Alpha, 59 multiple Delta, and 108 multiple Omicron variants. 60 flu and 60 RSV-positive samples were included as negative controls (total sample number=349). The conventional RADT showed sensitivity, specificity, positive predictive value (PPV), and negative predictive value (NPV) of 62.4% (95%CI: 54-70), 100% (95%CI: 97-100), 100% (95%CI: 100-100), and 58% (95%CI: 49-67) respectively. These measurements were enhanced using the Finecare™ RADT: sensitivity, specificity, PPV, and NPV were 92.6% (95%CI: 89.08-92.3), 96% (95%CI: 96-99.61), 98% (95%CI: 89-92.3), and 85% (95%CI: 96-99.6) respectively. The sensitivity of both RADTs could be greatly underestimated because nasopharyngeal swab samples collected UTM and stored at −80 °C were used. Despite that, our results indicate that the Finecare™ RADT is appropriate for clinical laboratory and community-based surveillance due to its high sensitivity and specificity.

## 1. Introduction

In 2021, the WHO has designated a new SARS-CoV-2 variant of concern and interest (VOC/I), including Delta (B.1.617.2), Lambda (C37), Mu (B.1.621), and Omicron (B.1.1.529) variant [1]. The emergence of novel SARS-CoV-2 variants has prompted concerns about increased infectivity, outbreaks among vaccinated people, and the feasibility of current test strategies [2]. Currently, quantitative real-time reverse transcription polymerase chain reaction (RT-qPCR) is considered the gold standard for SARS-CoV-2 diagnosis [3]. Despite its superior clinical performance, RT-qPCR is challenging to implement due to its expensive reagents, longer time to result, and requirements for sophisticated instrumentation [2-4].

Although not as sensitive as RT-qPCR, rapid antigen detection tests (RADTs) based on lateral flow immunoassay (LFIA) technology provide a fast, inexpensive, portable, and effective method of testing in the laboratory and non-laboratory settings [4]. RADTs have been successfully implemented to control HIV and malaria [5] and are quickly becoming an essential tool in SARS-CoV-2 testing [6]. The laboratory evaluations [7-9], and a Cochrane meta-analysis [10], have revealed a highly variable performance of RDATs, resulting in an ongoing debate over the utility of these tests for the detection of acute SARS-CoV-2-infection Several RADTs have already been approved for clinical use. According to WHO, it is recommended to have 80% sensitivity (true positive) and ≥ 97% specificity (true negative) for RADTs [11]. However, the emergence of the new variant has dramatically decreased the sensitivity of the RADT assays [12, 13]. Therefore, to increase the sensitivity of the conventional RADT assay for the detection of SARS-CoV-2 “N” antigen, Guangzhou Wondfo Biotech Co., Ltd. has developed a new Finecare™ RADT. The new Finecare™ RADT is an immunofluorescent-based assay, where the anti-N detecting antibodies are labeled or conjugated with fluorescent nanobeads. Finecare™ is a very sensitive meter used for the detection of the fluorescent signal and results interpretation of different RADT, qualitatively and quantitatively, including hemoglobin A1c (HBA1c), thyroid stimulating hormone (TSH), Beta-human chorionic gonadotropin (B-HCG), etc. However, the Wondfo conventional RADT is a colorimetric and qualitative assay where the anti-N detecting antibodies are labeled or conjugated with colloidal gold nanobeads. The test result of the conventional RADT is read out visually. The current study aimed to evaluate the performance of the conventional and the Fincecare RADT in detecting different SARS-CoV-2 variants.

## 2. Methods

### 2.1. Sample Collection

This study was conducted in full accordance with the research regulation at Hamad Medical Corporation (HMC) and Qatar University (QU). HMC-Institutional Review Board (HMC-IRB approval MRC-01-20-145) and QU-IRB (QU-IRB 1289-EA/20) reviewed and approved the study. The ethics committee waived the requirement of written informed consent for participation. Nasopharyngeal swabs suspended in universal transport media (UTM) from individuals confirmed with SARS-CoV-2 infection by RT-qPCR were received from the HMC virology lab between May 29, 2020, and January 22, 2022. The cycle threshold (CT) results of the RT-qPCR of each sample were also provided. Viral transport media were received at Qatar University Biomedical Research Center (QU-BRC) and subjected to sequencing, and then stored at −80°C. 187 UTM that are positive with different variants were selected for RADT, comprising 60 Alpha, 59 multiple Delta, and 108 multiple Omicron variants (see variant details in Table 3). A total of 60 flu and 60 RSV-positive samples were included as negative controls, making the overall sample total number 349.

### 2.2. Real-time reverse-transcription polymerase chain reaction testing

All PCR testing was conducted at the Hamad Medical Corporation (HMC) Central Laboratory, Sidra Medicine Laboratory, or National reference laboratory (NRL) following standardized protocols. Nasopharyngeal and/ or oropharyngeal swabs were collected from patients and placed in universal transport media (UTM). The RNA was extracted using KingFisher Flex (Thermo Fisher Scientific, USA), MGISP-960 (MGI, China), or ExiPrep 96 Lite (Bioneer, South Korea). The real-time reverse-transcription PCR (RT-qPCR) using TaqPath COVID-19 Combo Kits (Thermo Fisher Scientific, USA) on an ABI 7500 FAST (Thermo Fisher Scientific, USA) targeting the viral S, N, and ORF1ab gene regions, tested directly on the Cepheid GeneXpert system using the Xpert Xpress SARS-CoV-2 (Cepheid, USA) targeting the viral N and E gene regions, or loaded directly into a Rochecobas 6800 system and assayed with the cobas SARS-CoV-2 Test (Roche, Switzerland) targeting the ORF1ab and E-gene regions.

### 2.3. Sequencing: Determination of SARS-CoV-2 variants

The RNA was extracted from UTM using one of the previously described methods. The sequencing was performed at QU-BRC using the MGI G50 test, CleanPlex for MGI SARS-CoV-2. Research and Surveillance Panel (Paragon Genomics, USA), and DNBSEQ-G50RS High-throughput Sequencing Kit (FCL PE100/FCS PE150) were used according to the manufacturer’s protocols. Data analysis was conducted using a script, and FASTA fields were submitted to Bioinformatic Pangolin to identify the SARS-CoV-2 lineages. All SARS-CoV-2 samples were sequenced to determine SARS-CoV-2 VOC/Is, except for Alpha samples, there were not sequenced. Alpha samples were selected based on the collection date when the Alpha variant was the most prevalent between May and June 2020.

### 2.4. Conventional RADT

The conventional RADT (Cat No. W634; Wondfo, China) was performed according to the manufacturer’s instructions. In brief, the provided collection swab in the kit was embedded in the viral transport media then the swab was mixed with the provided extraction buffer tube in the kit. Three to four drops (80μl) were applied through a nozzle cap onto the test strip. The test strip (containing the gold beads labeled Ab) results were read out visually after 15 minutes. As recommended by the manufacturer’s reference guide, faint lines were considered positive if the control line was also present.

### 2.5. Finecare™ RADT

The Fincare™ RADT (Cat No: W286; Wondfo, China) was performed according to the manufacturer’s instructions, as exactly described above in section 2.3. Only the test strip (containing the fluorescent-labeled Ab) results were read after 15 minutes using the Finecare™ FIA Meter Plus (Wondfo, China). Finecare™ Meter Plus automatically calculates test results and presents them using the cut-off index (COI) unit. A COI greater than 1.0, was interpreted as positive; A COI less than 1.0, was interpreted as negative. The more SARS-COV-2 “N” antigen in the sample, the higher the signal value scanned by the Finecare™, and the greater the sample reading.

### 2.6. Statistical analysis

Data were analyzed using GraphPad Prism 9.3.1 (San Diego, CA, USA). An unpaired T-test was used to compare negative and positive results, and P-values ≤ 0.05 were considered statistically significant. One-way ANOVA was performed to compare the VOC/I RADT results, and P-values ≤ 0.05 were considered statistically significant. A correlation test between cycle threshold (CT) and Finecare™ RADT was performed in all graphs; the significance was * p ≤ 0.05, ** p ≤ 0.01, or *** p ≤ 0.001. Specificity and sensitivity with 95% confidence intervals and positive and negative predictive values of the RADTs were calculated using the RT-PCR as a reference method.

## 1. Results

### 1.1. Demographic and Clinical Information

Among the 349 samples, a total of 187 SARS-CoV-2 variant samples were included in the study, comprising 68 Omicron (median age: 35, IQR: 21-53), 59 Delta (median age: 33, IQR: 18-45), and 60 Alpha (median age: 36, IQR: 29-45) SARS-CoV-2 samples (Table S1). The negative controls included 60 flu (median age: 28, IQR: 9-38) and 60 RSV (median age: 5, IQR: 4-5) positive samples. Omicron lineages included BA.4 (n=20) and BA.5 (n=20) as they were the most prevalent Omicron lineages worldwide (Table S2).

### 1.2. Rapid Antigen Detection Tests (RADTs) enhanced by the Finecare™ RADT

Finecare™ RADT yielded more positive results (n=214, 66%) in comparison to conventional RADT (n=142, 41%) (Table 1). Moreover, the Finecare™ RADT showed higher sensitivity (92.6%, 95%CI: 89.08-92.3) in comparison to conventional RADT (62.4%, 95%CI: 54-70). Nevertheless, the conventional RADT showed higher specificity (100%, 95%CI: 100-100) in comparison to Finecare™ RADT (96%, 95%CI: 96-99.61). Moreover, conventional RADT showed higher PPV (100%, 95%CI: 100-100), but lower NPV (58%, 95%CI: 49-67) when compared to Finecare™ RADT (PPV=98%, 95%CI: 89-92.3; NPV=85%, 95%CI: 96-99.6).

**Table 1.** RADT results.

|  |  | **Conventional RADT** |  | **Finecare™ RADT** |  |
| --- | --- | --- | --- | --- | --- |
|  |  | **(n=347)** |  | **(n=326)** |  |
|  |  | Positive | Negative | Positive | Negative |
| **RSV (Conventional RADT n=60, Finecare™ RADT n=39)** |  | 0 (0%) | 60 (100%) | 3 (8%) | 36 (92%) |
| **Flu (Conventional RADT n=60, Finecare™ RADT n=60)** |  | 0 (0%) | 60 (100%) | 1 (2%) | 59 (98%) |
| **SARS-CoV-2** | *Omicron ( n=68) | 47 (69%) | 21 (30.8%) | 66 (97.1%) | 2 (2.9%) |
|  | Omicron BA.4 (n=20) | 14 (70%) | 6 (n=30) | 17 (85%) | 3 (15%) |
|  | Omicron BA.5 (n=20) | 14 (70%) | 6 (30%) | 19 (95%) | 1 (5%) |
|  | +Delta (n=59) | 30 (50.8%) | 29 (49.2%) | 59 (100%) | 0 (0%) |
|  | Alpha (n=60) | 37 (62%) | 23 (39%) | 49 (83%) | 11 (18.6%) |
| **Total** |  | 142 (41%) | 205 (59%) | 214 (66%) | 112 (34%) |
| **Sensitivity (95% CI)** |  | 62.4% (54-70) |  | 92.6% (89.08 – 92.3) |  |
| **Specificity (95% CI)** |  | 100% (100-100) |  | 96% (96- 99.61) |  |
| **PPV (95% CI)** |  | 100% (97-100) |  | 98% (89-92.3) |  |
| **NPV (95% CI)** |  | 58% (49-67) |  | 85% (96-99.6) |  |

\* B.1.1.529 (n=9), BA.1 (n=16), BA.2 (n=43, 39.8%); total 68. + See table 3 for detailed lineage classification.

The RADTs were then stratified by Ct values, and the sensitivity was investigated in relation to viral load. The sensitivity of both RADTs decreased when the Ct values increased. The conventional RADT showed a sensitivity of 75% for RT-PCR Ct values <20, moderate sensitivity of 57% for Ct values between 20 and 25, and sensitivity decreased dramatically at Ct values >25 (6%) (Table 2). Similarly, the Finecare™ RADT showed the highest sensitivity for RT-PCR Ct values <20 (98%), moderate sensitivity for Ct values between 20 and 25 (91%), and sensitivity decreased dramatically at CT values >25 (58.8%) (Table 2).

**Table 2.**
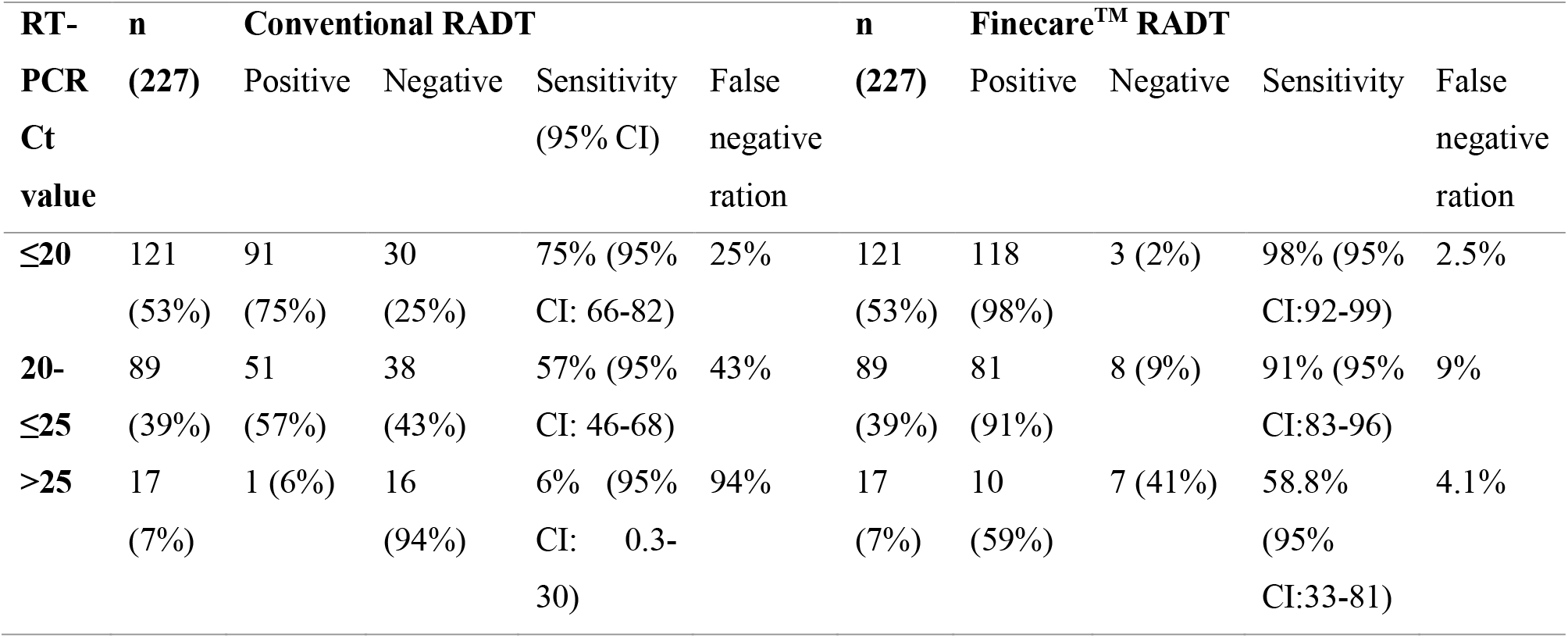
Sensitivity of the conventional RADT and Finecare™ RADT by reverse transcription polymerase chain reaction (RT-PCR) cycle threshold (Ct) intervals.

### 1.3. Detection of SARS-CoV-2 variants and lineages enhanced by the Finecare™ RADT

The performance of the conventional RADT and Finecare™ RADT in the detection of SARS-CoV-2 variants was evaluated (Figure 1). Overall, Finecare™ RADT showed higher sensitivity among all SARS-CoV-2 in comparison to the conventional RADT; nevertheless, conventional RADT showed slightly higher specificity (Figure 1).

**Figure 1.**
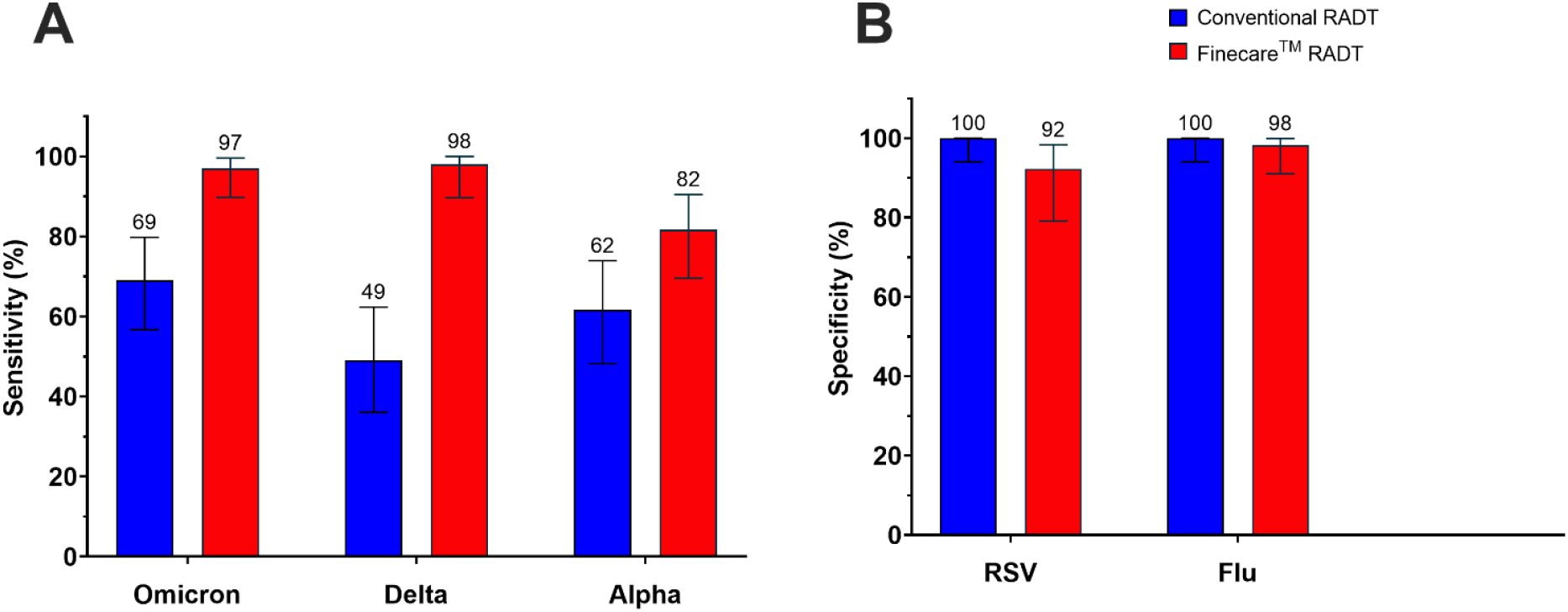
Analytical performance of Conventional and Finecare™ RADT, A) Sensitivity % among SARS-CoV-2 variants, and B) Specificity % among negative controls (RSV and Flu). Plotted values indicate the % sensitivity and % specificity. Horizontal black bars indicate the confidence intervals (CIs).

A detailed assessment of the performance of the conventional RADT and the Finecare™ RADT across SARS-CoV-2 lineages was performed (Table 3). Overall, Finecare™ RADT detection of SARS-CoV-2 lineages was higher (92.1%, n=209 positives) in comparison to the conventional RADT (63%, n=142 positives) (Table 3). The conventional RADT detected all Delta SARS-CoV-2 lineages, excluding the AY.106, AY.114, AY.120, AY.34, and AY.5.2. With the exception of the AY.34 lineage, the Finecare™ RADT detected all Delta SARS-CoV-2 lineages.

**Table 3.** Conventional RADT and Finecare™ RADT detailed test results of SARS-CoV-2 lineages

| Lineage/RADT test | Conventional RADT |  | Finecare™ RADT |  |
| --- | --- | --- | --- | --- |
|  | Positive | Negative | Positive | Negative |
| **Omicron (n=108)** | 76 (70%) | 32 (30%) | 102 (94%) | 6 (6%) |
| **B.1.1.529 (n=9, 8.3%)** | 5 (4.6%) | 4 (3.7%) | 9 (8.3%) | 0 (0%) |
| **BA.1 (n=16, 14.8%)** | 8 (7.4%) | 8 (7.4%) | 14 (13%) | 2 (1.9%) |
| **BA.2 (n=43, 39.8%)** | 35 (32.4%) | 8 (7.4%) | 43 (39.8%) | 0 (0%) |
| **BA.4 (n=12, 11.1%)** | 8 (7.4%) | 4 (3.7%) | 10 (9.3%) | 2 (1.9%) |
| **BA.4.1 (n=6, 5.6%)** | 5 (4.6%) | 1 (0.9%) | 5 (4.6%) | 1 (0.9%) |
| **BA.4.5 (n=2, 1.9%)** | 1 (0.9%) | 1 (0.9%) | 2 (1.9%) | 0 (0%) |
| **BA.5.1 (n=3, 2.8%)** | 2 (1.9%) | 1 (0.9%) | 3 (2.8%) | 0 (0%) |
| **BA.5.2 (n=14, 13%)** | 9 (8.3%) | 5 (4.6%) | 13 (12%) | 1 (0.9%) |
| **BA.5.2.1 (n=2, 1.9%)** | 2 (1.9%) | 0 (0%) | 2 (1.9%) | 0 (0%) |
| **BA.5.3.1 (n=1, 0.9%)** | 1 (0.9%) | 0 (0%) | 1 (0.9%) | 0 (0%) |
| **Delta (n=59)** | 29 (49.2%) | 30 (50.8%) | 58 (98%) | 1 (2%) |
| **AY.102 (n=1, 1.7%)** | 1 (1.7%) | 0 (0%) | 1 (1.7%) | 0 (0%) |
| **AY.106 (n=1, 1.7%)** | 0 (0%) | 1 (1.7%) | 1 (1.7%) | 0 (0%) |
| **AY.114 (n=1, 1.7%)** | 0 (0%) | 1 (1.7%) | 1 (1.7%) | 0 (0%) |
| **AY.116 (n=1, 1.7%)** | 1 (1.7%) | 0 (0%) | 1 (1.7%) | 0 (0%) |
| **AY.120 (n=1, 1.7%)** | 0 (0%) | 1 (1.7%) | 1 (1.7%) | 0 (0%) |
| **AY.122.1 (n=1, 1.7%)** | 1 (1.7%) | 0 (0%) | 1 (1.7%) | 0 (0%) |
| **AY.26 (n=1, 1.7%)** | 1 (1.7%) | 0 (0%) | 1 (1.7%) | 0 (0%) |
| **AY.32 (n=1, 1.7%)** | 1 (1.7%) | 0 (0%) | 1 (1.7%) | 0 (0%) |
| **AY.33 (n=2, 3.4%)** | 2 (3.4%) | 0 (0%) | 2 (3.4%) | 0 (0%) |
|  | Positive | Negative | Positive | Negative |
| **AY.34 (n=1, 1.7%)** | 0 (0%) | 1 (1.7%) | 0 (0%) | 1 (1.7%) |
| **AY.4 (n=13, 22%)** | 5 (8.5%) | 8 (13.5%) | 13 (22%) | 0 (0%) |
| **AY.43 (n=2, 3.3%)** | 1 (1.7%) | 1 (1.7%) | 2 (3.4%) | 0 (0%) |
| **AY.47 (n=1, 1.7%)** | 1 (1.7%) | 0 (0%) | 1 (1.7%) | 0 (0%) |
| **AY.5 (n=1, 1.7%)** | 1 (1.7%) | 0 (0%) | 1 (1.7%) | 0 (0%) |
| **AY.5.2 (n=1, 1.7%)** | 0 (0%) | 1 (1.7%) | 1 (1.7%) | 0 (0%) |
| **AY.9 (n=1, 1.7%)** | 1 (1.7%) | 0 (0%) | 1 (1.7%) | 0 (0%) |
| **B.1.617.2 (n=29, 49.2%)** | 13 (22%) | 16 (27%) | 29 (49.2%) | 0 (0%) |
| **Alpha (n=60)** | 37 (62%) | 23 (38%) | 49 (82%) | 11 (18%) |

As the Ct value is inversely related to the viral load, the RT-qPCR Ct value was investigated across SARS-CoV-2 variants using the conventional RADT and Finecare™ RADT (Figure 2) [14]. Ct values were significantly higher (p≤ 0.001) among individuals with false-negative conventional RADT compared to true positives across Omicron samples. Similarly, Ct values were significantly higher (p≤ 0.05) among individuals with false-negative Finecare™ RADT compared to true positives across Omicron samples. In relation to the Alpha sample, Ct values were significantly higher (p ≤ 0.0001) among individuals with a false-negative conventional RADT compared to true positives across Omicron samples. Similar results were obtained from Finecare™ RADT (p≤ 0.001). On the contrary, Delta samples didn’t show any significance using both the conventional RADT and Finecare™ RADT.

**Figure 2.**
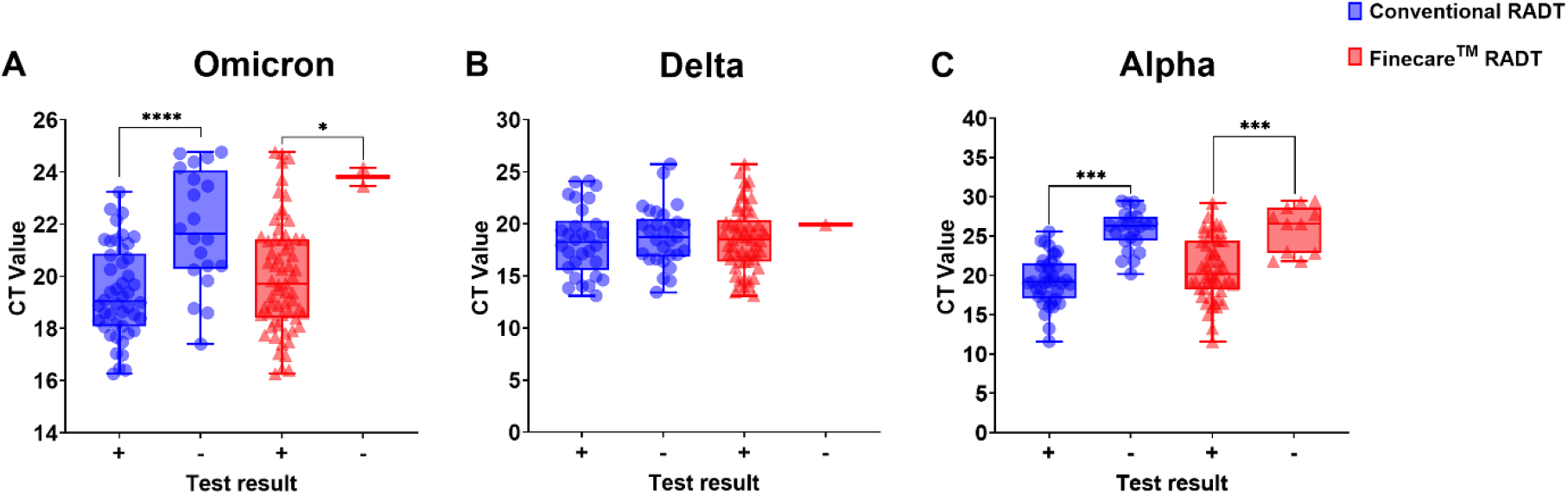
Box blot representing RADT among SARS-CoV-2 variant against Ct value, A) Omicron, B) Delta, and C) Alpha. Significant level * p ≤ 0.05, ** p ≤ 0.01, or *** p ≤ 0.001.

### 1.4. Rapid antigen detection tests (RADT) performance in the detection of Omicron BA.4 and BA.5 lineages

Furthermore, we evaluated the performance of RADTs in the diagnosis of BA.4 (n=20) and BA.5 (n=20) lineages as they harbor unique mutations, including changes referred to as L452R and F486V in the viral spike protein that may weaken their ability to latch onto host cells (Table S2). Ct values showed no significant difference among individuals with false-negative compared to true positives in both conventional RADT test and Finecare™ RADT across BA.4 and BA.5 samples (Figure 3).

**Figure 3.**
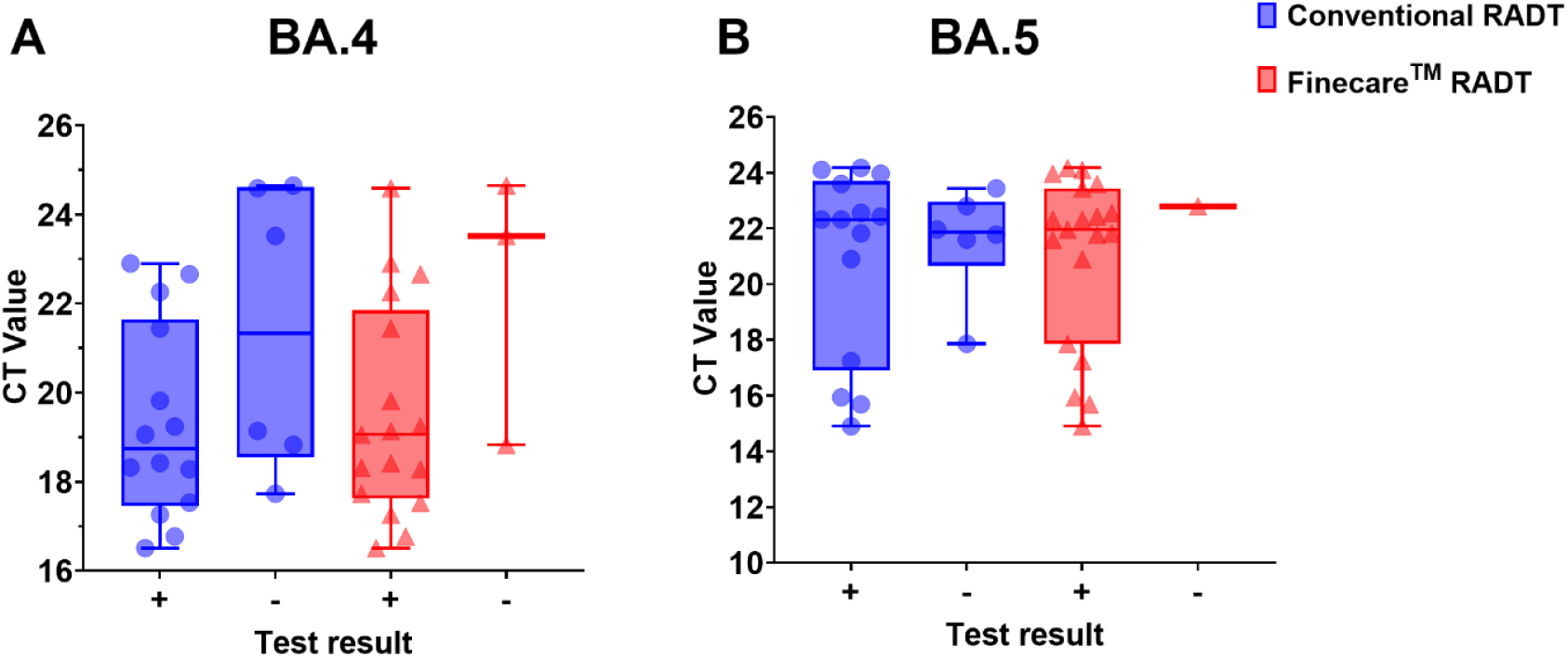
Box blot representing RADT among SARS-CoV-2 variant against Ct value, A) BA.4, and B) BA.5. Significant level * p ≤ 0.05, ** p ≤ 0.01, *** p ≤ 0.001.

## 2. Discussion

Rapid identification of SARS-CoV-2-infected individuals is essential to limit viral transmission [15]. So far, several RDATs have been developed by various manufacturers from multiple countries, mainly based on either colloidal gold- or immunofluorescence-based assays. Nevertheless, a comparative approach investigating the efficacy of both types of RDATs, and whether they can be relied on for SARS-CoV-2 diagnosis, particularly in the context of the newly emerged SARS-CoV-2 variants, is lacking. In the current study, we evaluated the performance of Finecare™ RADT for the detection of different SARS-CoV-2 variants using the RT-PCR test results as a reference test.

The Finecare™ RADT showed an overall better performance compared to conventional RADT (Table 1, Figure 1) in detecting Omicron, Delta, and Alpha SARS-CoV-2 variants, with an overall sensitivity of 92.6%, and a PPV of 92.6%, in comparison to the conventional RADT (sensitivity: 62.4%, PPV: 62.4%). Nevertheless, the conventional RADT showed higher specificity (specificity: 100%, NPV: 100%) compared to Finecare™ RADT (specificity: 96%, NPV: 96%) (Table 1). According to the WHO, the accepted sensitivity and specificity are ≥ 80% and ≥ 97%, respectively, while the European Center for Disease Prevention and Control recommends ≥ 90% sensitivity and ≥ 97% specificity [16, 17]. This has been a challenging task because almost all of the currently available RDATs were reported to have sensitivity disadvantages. For instance, a recent meta-analysis evaluated the overall performance of immunofluorescent-based and colloidal gold-based-based RDATs and reported an overall high pooled specificity of ∼99.4%, but poor sensitivity of ∼68.4% [10]. Similarly, a systematic review conducted by Lee *et al*., revealed an overall sensitivity of 68% [18]. Furthermore, another review summarized the accuracy of multiple RDATs, reporting an average sensitivity of 72.0% in symptomatic individuals [10]. Thus, compared to other RDATs, Finecare™ RADT seems to be top-performing in terms of achieving high sensitivity (92.6%) and high specificity (96%) (Table 1) and thus could serve as a promising alternative to RT-PCR for SARS-CoV-2 diagnosis. It is noteworthy to mention that the sensitivity of both RADTs could be greatly underestimated because nasopharyngeal swab samples were collected in UTM which diluted the sample. Also, for both RADTs, the nasopharyngeal swab samples collected in UTM where further diulted in to the RADTs buffer. Rather than directly performing the test from the patient nasopharyngeal swab. Beside the fact that some smple were sore in the −80°C for more than a year.

We further investigated the sensitivity in relation to the Ct-value. The sensitivity of both Finecare™ RADT decreased dramatically with increasing Ct-value (decreasing viral load) (Table 2, Figure 2). For Ct-values <20, 20 - 25, and >25, the conventional RADT showed sensitivities of 75%, 57%, and 6%, respectively, while Finecare™ RADT showed sensitives of 96%, 91%, and 58.8%, respectively (Table 2, Figure 2). False-negative results were observed across high CT values (low viral load) (Table 2). These findings are in concordance with other studies where a negative correlation of Ct values of RT-PCR and RADTs sensitivity was identified [18]. Currently, there is no definitive Ct-value threshold beyond which antigen tests consistently yield false-negative results. However, a recent study revealed that RDATs are frequently negative in PCR-positive samples with Ct-values above 24–28 [19]. Similarly, another study comprising 468 samples from a German maximum care hospital identified Ct-values ≤ 22 as the limit for a 100% correlation of PCR and RADT [20]. Based on these findings, early detection using RDATs kits is recommended. Nevertheless, according to the Centers for Disease Control and Prevention (CDC), a higher Ct value (>33) reflects a non-contagious stage [21], which justifies the use of RDATs kits.

Except for a few Delta lineages, such as AY.106, AY.114, AY.120, AY.34, and AY.5.2, the conventional RADT was able to detect all SARS-CoV-2 variants. Similarly, the Finecare™ RADT detected all SARS-CoV-2 variants except for the AY.34 Delta lineage. Nevertheless, the Finecare™ RADT detected higher SARS-CoV-2 variants than the conventional RADT. That is due to the fact Finecare™ RADT is coupled with an FIA meter. This lowers the limit of detection in comparison to other common RADTs [22]. Moreover, both RADTs successfully detect circulating SARS-CoV-2 VOC/I BA.4 and BA.5 (Table 3, Figure 4).

This study had some limitations. First, our findings were limited to circulating SARS-CoV2 lineages at the time of the investigation. Alpha specimens were not sent for NGS; they were assumed to be Alpha based on the swab collection date. As the clinical manifestations were missing, no distinction was made between symptomatic and non-symptomatic participants, which may influence the sensitivity of this rapid antigen assay. After RT-PCR, samples were frozen in UTM and examined for this study after a period of time that might lead to antigen dilution and degradation.

## 3. Conclusion

In conclusion, RADTs successfully detected all SARS-CoV-2 variants with high viral loads. Due to the test’s high sensitivity and specificity, the immunofluorescence-based Finecare™ RADT is suitable for both clinical and community-based surveillance. Moreover, considering the short turnaround time, user-friendliness, and low costs we believe that in the context of community-based surveillance of symptomatic individuals, these advantages outweigh the lower sensitivity of Wonfo 2019-nCoV-2 Antigen test compared to RT-qPCR.

## Data Availability

All data produced in the present work are contained in the manuscript

## Data availability

All data produced in the present study are available upon reasonable request to the authors.

## Authors’ contributions

Conceptualization: GKN, HMY. Participant recruitment and demographic data collection: GKN, HMY, FA, HA. Laboratory testing: FA. Supervision: HMY, GKN. Data analysis: FA, SY, GKN. First draft writing: FA, SY. Review and editing: GKN, HMY.

## Acknowledgment

We thank the many dedicated persons at Hamad Medical Corporation, Sidra Medicine, and the National Reference lab for their diligent efforts and contributions to making this study possible.

