## Supplemental Table 1 and Table 2 for "Enhancing the sensitivity of rapid antigen detection test (RADT) of different SARS-CoV-2 variants and lineages using fluorescence-labeled antibodies and a fluorescent meter"

**Table S1.** Demographic Data of selected samples.

| Sample | Omicron (n=68) | Delta (n=59) | Alpha (n=60) | Flu (n-60) | RSV (n=60) |
|---|---|---|---|---|---|
| **Median age in years (IQR)** | 35 (21-53) | 33 (18-45) | 36 (29-45) | 28 (9-38) | 5 (4-5) |
| **Gender** | | | | | |
| **Male** | 37 (53.6%) | 31 (51.7%) | 45 (75%) | 30 (50%) | 30 (50%) |
| **Female** | 32 (46.4%) | 29 (48.3%) | 15 (25%) | 30 (50%) | 30 (50%) |
| **Region** | | | | | |
| **MENA** | 31 (44.9%) | 28 (46.7%) | 25 (41.7%) | | 51 (85%) |
| **Non-MENA** | 38 (55.1%) | 32 (53.3%) | 35 (58%) | | 9 (15%) |
| **Not provided** | | | | 60 (100%) | |

Table S2. Demographic Data of Omicron BA.4 and BA.5 lineages

Commented [GKN1]: Suplment

Commented [SY2R1]: done

| **Omicron Lineage** | **BA.4** **(n=20)** | **BA.5** **(n=20)** |
|---|---|---|
| **Median age in years (IQR)** | 42 (26-45) | 31 (17-50) |
| **Gender** | | |
| **Male** | 8 (40%) | 10 (50%) |
| **Female** | 12 (60%) | 10 (50%) |
| **Region** | | |
| **MENA** | 9 (45%) | 10 (50%) |
| **Non-MENA** | 11 (55%) | 10 (50%) |
